# Antibiotic courses for acute otitis media alter human gut microbiome resistome and colonization resistance against antimicrobial resistant species in children

**DOI:** 10.1101/2022.04.08.22273642

**Authors:** Weizhong Li, Katja Kielenniva, Claire Kuelbs, Mark Novotny, Tero Kontiokari, Suvi Sarlin, Mysore Tejesvi, Karen E. Nelson, Terhi Tapiainen

## Abstract

Antimicrobial resistance (AMR) is a major global public health problem. Human gut microbiome plays an important role in modulating AMR. On one hand, the microbiome itself can serve as a reservoir of AMR genes, i.e. resistome. On the other hand, the microbiome performs colonization resistance, preventing invasive microbes from colonizing the gastrointestinal tract. In this study, we investigated how antibiotic treatment affects the resistome and colonization resistance of the gut microbiome in children receiving amoxicillin, amoxicillin-clavulanate, or no treatment for acute otitis media in a randomized clinical trial. Fecal samples from children receiving an antibiotic or no treatment before and after the treatment were analyzed using deep metagenomic sequencing. We used a flow cytometry-based approach to quantify the bacterial load in the fecal samples. Both metagenomic sequencing-based relative abundance and flow cytometry-based absolute abundance of the microbial species were analyzed. We found that the resistome fluctuated over time and in a small fraction (∼10%) of subjects, AMR genes increased rapidly due to colonization by AMR species, even in the control group without any antibiotic treatment. Amoxicillin significantly increased the risk for invasive species, especially pathogenic species carrying AMR genes, to colonize the gut. We also found that children lacking *Blautia, Ruminococcus, Faecalibacterium, Roseburia*, or *Faecalitalea* were more vulnerable to colonization by invasive AMR species in their gut microbiome.

## Introduction

Antimicrobial resistance (AMR) is a major global public health problem. According to the Centers for Disease Control and Prevention (CDC), at least 2 million people in the United States acquire serious infection with AMR bacteria and among them at least 23,000 die annually^1^. Over the last 15 years, research on the human microbiome has allowed the scientific community to gain unprecedented knowledge on the diversity, composition, functions and dynamics of human microbiome communities at different body sites^2–5^. Research by various groups, including our own, has revealed the complex interactions between the microbiome, pathogenic species and different antibiotics^6–11^. Antibiotics have a major and prolonged impact on the human microbiome^12^. Since the human microbiome plays important roles in preventing colonization of pathogenic microbes, educating the immune system, and producing metabolites that are important for various parts of the human body, the use of antibiotics can compromise the ability of the microbiome to protect its human host. This may be even more significant for young children, since the first years of life are the most important stage for the establishment of their microbiomes.

Young children frequently receive antibiotic prescriptions for common respiratory and other infections and many spend extensive periods of time in day care centers which increases the risk of otitis media, the most common reason for antibiotic treatment in children^13,14^. In epidemiological studies, antibiotic use has been linked with subsequent overweight^15^, asthma^16^, juvenile arthritis^17^, and inflammatory bowel disease^18^ in children. Observational studies of the intestinal microbiome in infants and young children have suggested that macrolides, including azithromycin, have prolonged effects on the gut microbiome^19^ whereas the impact of commonly used penicillin derivatives may be less clear. Epidemiological data suggests that the use of antimicrobials in children is closely connected with the AMR in children.^20,21^ Our recent microbiome study demonstrated the impact of antibiotic treatment given during labor or immediately after birth on the intestinal microbiome and the emergence of AMR genes and the main source of colonization^10,11,22^. Currently, there is a knowledge gap regarding the impact of different antibiotic agents on the intestinal microbiome in young children because controlled, randomized studies are lacking.

Here, we used non-severe acute otitis media as a clinical model to investigate human gut microbiome resistome and colonization resistance species in children. This enabled us to conduct a randomized trial in an ethically acceptable manner since the current clinical guidelines for treating non-severe otitis media in children allow both antibiotics treatment and watchful waiting without antibiotics as treatment options^23^. In the present study, the children were randomly allocated to receive either amoxicillin, amoxicillin-clavulanate, or no treatment, and children with previous amoxicillin allergy received azithromycin. We investigated how antibiotic treatment affected the resistome and colonization resistance of microbiome in these children through metagenomic sequencing of the fecal samples collected before and after the treatment. Furthermore, as studies have shown that bacteria load in the human gut varies between people and fluctuates significantly over time^24^, we quantified absolute bacterial load by a flow cytometry-based approach. In this study, both relative abundance and absolute abundance of the microbial species were analyzed.

## Results and discussion

### Study cohort

The study cohort included 62 Finnish children, 6 month to 6 years of age, and approximately 50% male and 50% female. All children had non-severe otitis media. They were randomly allocated to treatment groups receiving different antibiotics for 7 days and a control group receiving placebo. In the treatment group of 52 children, amoxicillin, amoxicillin-clavulanate and azithromycin were given to 25, 23 and 4 children respectively. Only the children who were allergic to amoxicillin received azithromycin. Children who switched to antibiotics treatment from the no treatment group due to treatment failure were analyzed in the antibiotics groups. The control group had 10 children who did not receive any antibiotics during the study. Stool samples before the antibiotic treatment (day 0) and after (day 10) were collected and analyzed (**Table S1**,**S2**).

In this study, we did analysis and comparison for the 4 individual groups (amoxicillin, amoxicillin-clavulanate, azithromycin and control), and also for the combined antibiotic treatment groups and the control group. In addition, the samples were divided into a baseline group without antibiotic disclosure (n=72, day 0 from all children and day 10 from the control group) and an affected group (n=52, day 10 from antibiotic treatment).

### Antibiotics have small effect on total bacterial load, but marked effect on microbial diversity

The bacterial load or absolute abundance, number of cells per gram fecal specimen, measured by flow-cytometry for these samples varied from 1.9×10^9^ to 4.8×10^10^, with the median, mean and standard deviation being 1.3×10^10^, 1.5×10^10^ and 9.8×10^9^ respectively (**Table S2**). This is within the range of the previously reported bacterial load such as the study by Vandeputte et al^24^ and the study by Galazzo et al^25^, which reported median per-gram cell count as 1.5×10^11^ and 2.3×10^10^ respectively. Our cell count values were slightly lower than those in these two previous studies, which is likely explained by the study population here including young children (mean age 42 months), while the two previous studies were for adult stool samples.

Antibiotic treatment appeared to have little effect on the bacterial load in gut microbiome in these children, as no significant difference was found between before and after the treatment (paired Wilcoxon rank sum test) in different antibiotic treatment groups (**Figure S1**). No significant difference was found either between the baseline and the affected samples (Wilcoxon rank sum test). Notable decrease in bacterial load was only observed in the children treated by azithromycin. There were only 4 children in this group, so it is difficult to assess the statistical significance (p = 0.1, paired Wilcoxon rank sum test). It is well acknowledged that azithromycin has a large impact on the gut microbiome^19^ which is in line with our observation showing that azithromycin reduced the bacterial load in the gut microbiome.

Although antibiotics didn’t impact the total bacterial load, the microbiome diversity was influenced by antibiotics. The relative taxonomy abundance at phylum, genus and species level are shown in **Table S3-S5**. The relative species abundance was converted into absolute abundance (**Table S6**) using the bacterial load data (see Methods). We calculated the alpha diversity of the samples, as the Shannon diversity index, using the absolute species abundance (**Table S7**). The Shannon diversity index in the antibiotics group was reduced after the treatment (p = 0.005, paired Wilcoxon rank sum test) (**Figure S2**). This significant reduction was not observed in the control group (p = 0.6, paired Wilcoxon rank sum test). Also the Shannon diversity index of the affected group is significantly lower than the baseline group (p=0.0007, Wilcoxon rank sum test). The beta diversity, calculated as the Bray-Curtis distance based on absolute species abundance, among the affected samples was significantly larger than those among the baseline samples with p-value of 1e-49 (Wilcoxon rank sum test). Analysis of alpha and beta diversity using the relative species abundance (**Table S8**), we got very similar results as using the absolute species abundance (data now shown).

### Antibiotics permitted more invasive species to enter and colonize the gut

By comparing the taxonomy abundance at the genus level before and after antibiotic treatment, several genera were found to be increased after the treatment. We used the paired Wilcoxon rank sum test on the absolute abundance for these genera. Then the raw p values were adjusted for multiple testing using the Benjamini & Hochberg (BH) method^26^ for false discovery rate (FDR). The results are shown in **Table S9**. *Klebsiella* is significantly increased (p = 0.0003, fdr=0.02) by antibiotics treatment from mean abundance of 3.5×10^6^ to 228.4×10^6^ cells/g, a 65 fold increase (**Figure 1a**). The abundance of *Citrobacter, Dialister, Enterococcus, Raoultella, Prevotella, Phascolarctobacterium, Sanguibacteroides, Yersinia, Kluyvera* and *Mediterranea* also increased (p < 0.05), although the differences are not statistically significant after multiple testing correction (fdr > 0.05) (**Figure 1a**). The antibiotic treatment caused the depletion of more taxa (**Figure 1b**). But the magnitude of the decrease is modest, usually just a few fold of decrease (**Table S9**). For the most common and abundant taxa found in these children, e.g. *Bifidobacterium, Blautia, Ruminococcus, Streptococcus, Clostridium, Collinsella, Mediterraneibacter, Dorea* and *Bacteroides*, antibiotics didn’t cause significant change in their abundance (p > 0.05) (**Table S9**).

**Figure 1.**
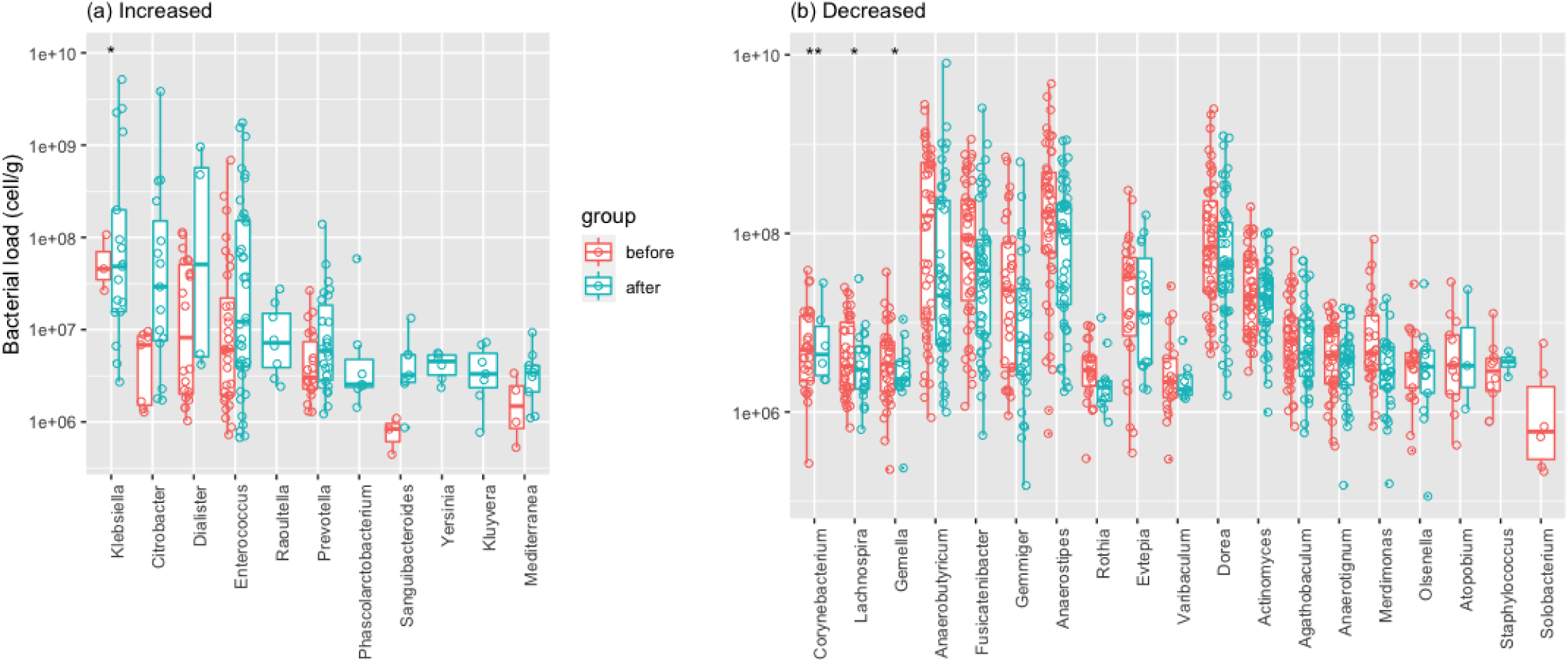
Boxplot of bacterial genera mostly impacted by antibiotic treatment. Y-axis is the bacterial load. Statistical significance annotation symbol: * P ≤ 0.05, ** P ≤ 0.01.

Using the same approach to compare the samples between the baseline and the affected groups, we obtained very similar results about the taxa change influenced by antibiotic treatment (**Figure S3, Table S10**).

### Antibiotics caused changes in gut microbiome resistome

The AMR genes in the samples were identified using RGI^27^ (see Methods). **Table S11** lists the abundance of AMR genes, in copy per million (CPM), which is the number of genes belonging to this AMR gene family per million genes found in the sample. The reported AMR genes in the CARD database ^27^ are organized into AMR gene families, AMR drug classes and AMR machemisms. Therefore, we also calculated the abundance for each AMR gene family (**Table S12**), AMR drug class (**Table S13**) and AMR machemism (**Table S14**).

We first analyzed the total AMR gene abundance of these samples. The antibiotic treatment caused a significant increase of AMR genes in these samples (**Figure 2**). In the amoxicillin group, the total AMR genes per sample were increased from 550 CPM before treatment to 2,918 CPM after treatment, ∼ 5 folds of increase (p = 0.05, paired Wilcoxon rank sum test). In the amoxicillin-clavulanate group, the total AMR genes per sample were increased from 593 CPM to 831 CPM (p=0.07, paired Wilcoxon rank sum test). No change was observed in the azithromycin group (p = 1.0, paired Wilcoxon rank sum test). Also in the control group, there was no significant change in total AMR gene abundance (p = 0.3, paired Wilcoxon rank sum test). When the three antibiotic treatment groups were combined in the analysis, we could observe a higher statistical significance (p = 0.01, paired Wilcoxon rank sum test) between before and after treatment samples (**Figure 2b**). Also, when the baseline samples and the affected samples were compared, more significant impact was observed that antibiotic treatment induced higher AMR genes in the microbiome (p = 0.005, Wilcoxon rank sum test) (**Figure 2c**).

**Figure 2.**
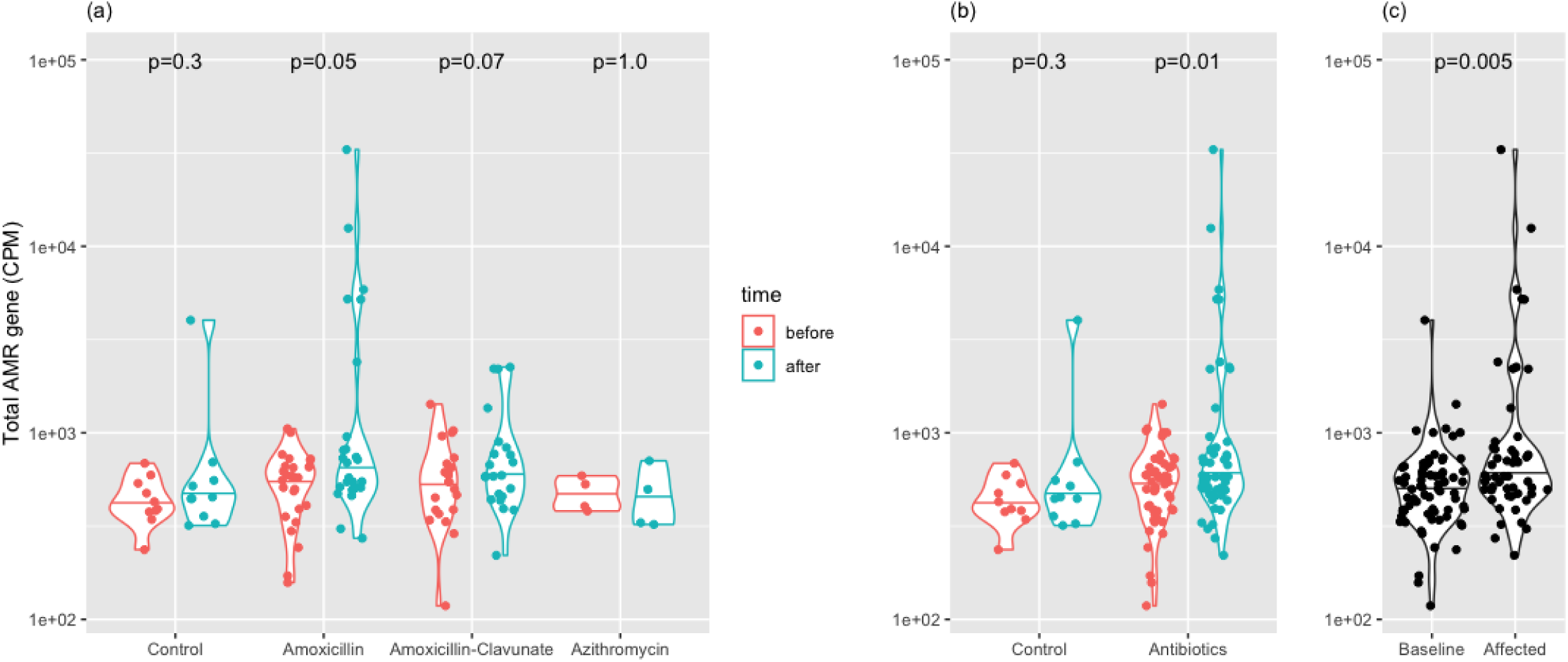
Total AMR gene abundance in fecal specimens before and after antibiotic treatments in the treatment and control groups (a), (b) and between the baseline and affected groups (c). The AMR gene abundance is expressed in copy per million (CPM), which is the number of AMR genes per million genes found in the sample.

Among the three different antibiotics, amoxicillin appeared to have the largest risk to allow the increase of AMR genes in the microbiome. This is possibly due to the fact that amoxicillin is the most prescribed antibiotic ^28^ and many bacteria have developed AMR against it. So, while amoxicillin was inhibiting many of the existing members of species in the native microbiome community, more invasive species carrying AMR against amoxicillin could still colonize the gut. Our data suggested that azithromycin may be able to prevent the colonization of AMR-carrying invasive species (**Figure 2a**), but this is not conclusive due to the small group size (N=4).

Next, we investigated which specific AMR genes were among the most increased. As azithromycin didn’t change the AMR gene abundance, we only compared the AMR genes between before and after treatment in amoxicillin and amoxicillin-clavulanate groups. AMR genes in 17 drug classes were significantly increased (p<0.05 and FDR < 0.05, paired Wilcoxon rank sum test) (**Figure 3, Table S15**). These AMR drug classes, in order of increasing p value, were cephamycin, peptide antibiotic, nitrofuran antibiotic, glycylcycline, benzalkonium chloride, elfamycin antibiotic, rhodamine, cephalosporin, rifamycin antibiotic, fluoroquinolone antibiotic, aminoglycoside antibiotic, penem, nitroimidazole antibiotic, triclosan, penam, mupirocin and diaminopyrimidine antibiotic. AMR genes in most of these drug classes exhibit over 10 fold of increase after antibiotic treatment (**Figure 3, Table S15)**. Amoxicillin appeared to lead to a higher abundance of AMR genes than amoxicillin-clavulanate (**Figure 3**). The children in the present study only received amoxicillin or amoxicillin-clavulanate, but AMR genes in multiple drug classes increased after the treatment.

**Figure 3.**
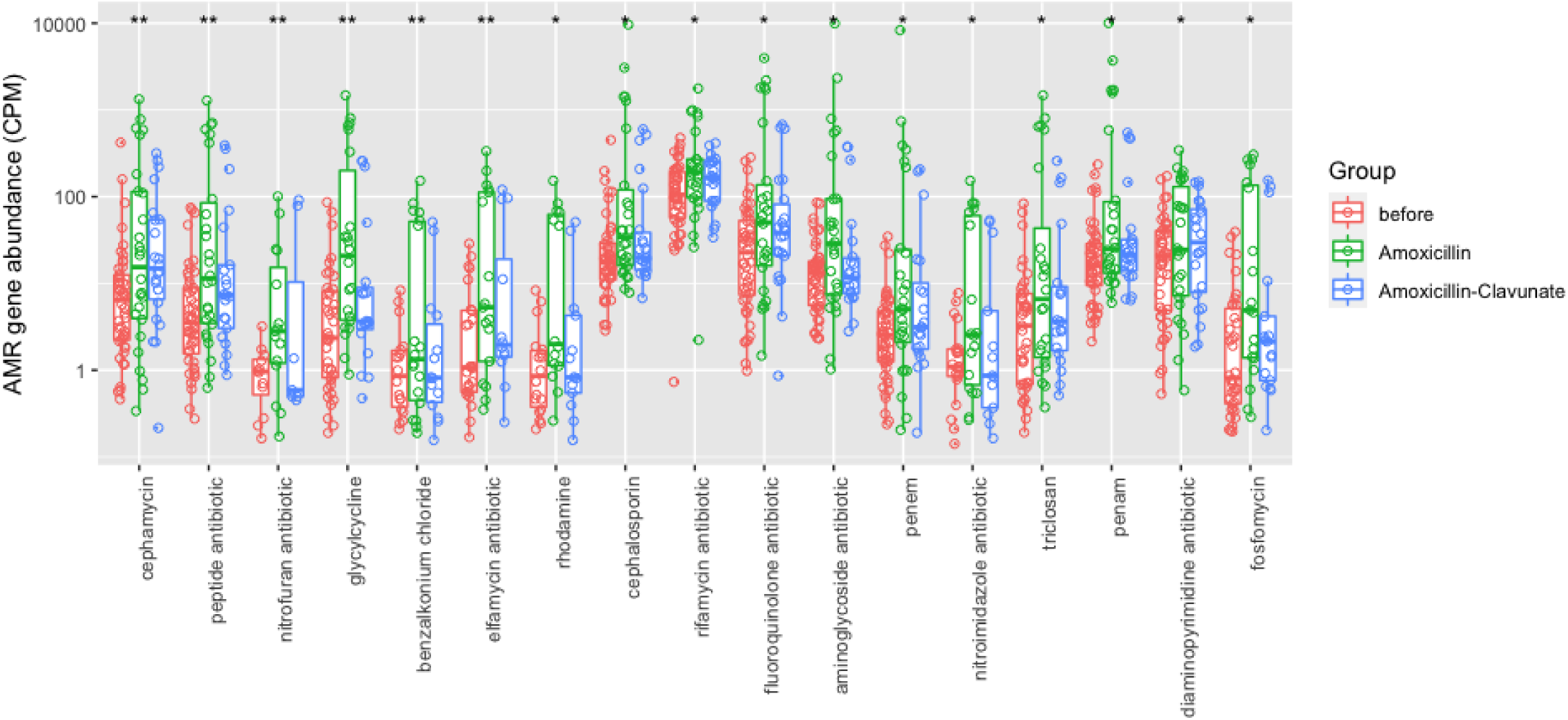
Significant AMR gene in drug classes between before and after amoxicillin and amoxicillin-clavulanate treatments. The AMR gene abundance in the y-axis is expressed in copy per million (CPM), which is the number of AMR genes per million genes found in the sample. Legend groups: before - samples before the treatment in amoxicillin and amoxicillin-clavulanate groups; amoxicillin - samples after treatment in amoxicillin group; amoxicillin-clavulanate - samples after treatment in amoxicillin-clavulanate group. Statistical significance annotation symbol: * P ≤ 0.05, ** P ≤ 0.01.

The bacterial species that carried the most AMR genes are listed in **Table 1** (full list in **Table S16**). These included species in genera *Escherichia, Klebsiella, Citrobacter, Shigella, Bifidobacterium* and *Enterobacter*. For example, AMR gene carrying *E. coli* were detected in 78 out of 124 samples. In one sample after amoxicillin treatment, AMR gene abundance contributed by *E. coli* reached 22,961 CPM. Most of these top AMR gene carrying species were potential pathogenic strains. We found several *Bifidobacterium* species, the core members in the normal human gut microbiome in these children, that carried AMR genes as well.

**Table 1.**
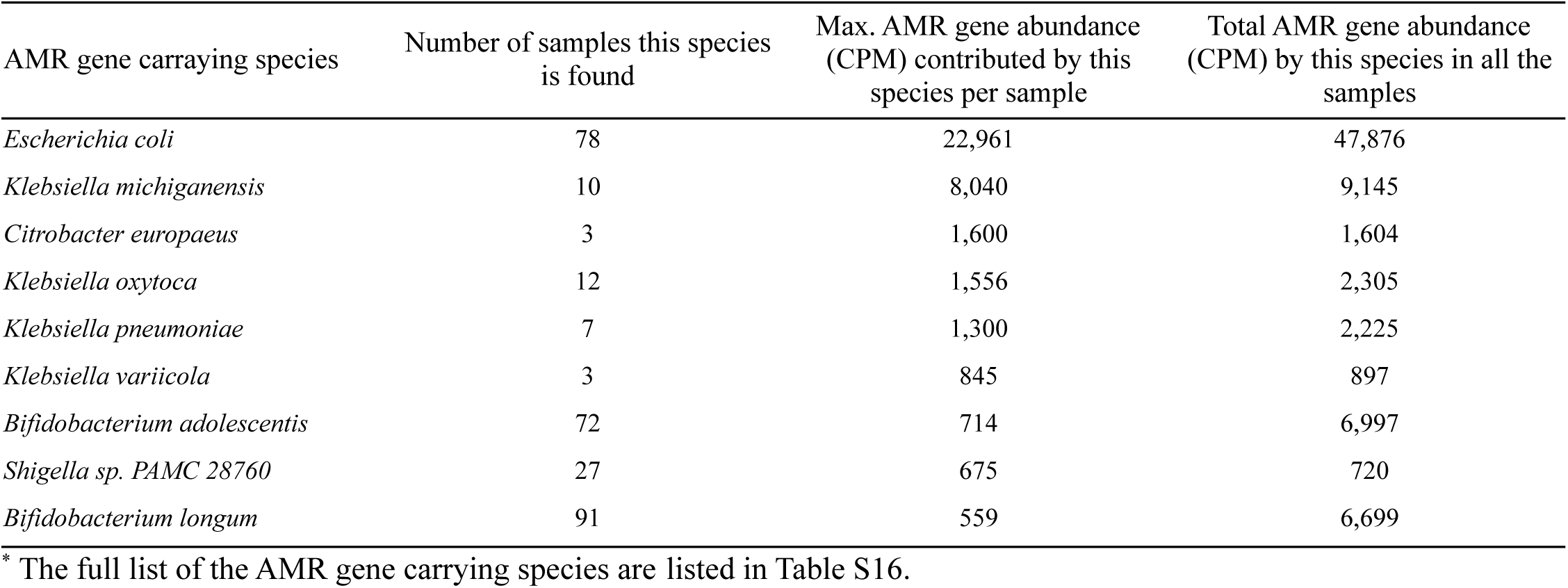
Top AMR gene carrying specie^*^

The composition and abundance of the AMR genes are shown in **Figure 4**. In the baseline samples (samples before the treatment and all the control samples), the AMR genes are mainly from *Bifidobacterium* and other taxa making the normal microbiome (**Figure 4**). Significant spike of AMR genes was seen in 6 children after treatment by amoxicillin (subjects: 15, 18, 25, 53, 54, 63, **Figure 4**) due to *E. coli, K. michiganensis* and *Klebsiella pneumoniae*. In the amoxicillin-clavulanate group, 3 children exhibited large increase in AMR genes (subjects: 2, 5, 56), owing to *E. coli, Citrobacter europaeus, Klebsiella oxytoca and Klebsiella pneumoniae* (**Figure 4**). In the control group receiving no antibiotic treatments, one sample (16B) were found to have elevated AMR genes, mostly contributed by *E. coli* (**Figure 4**).

**Figure 4.**
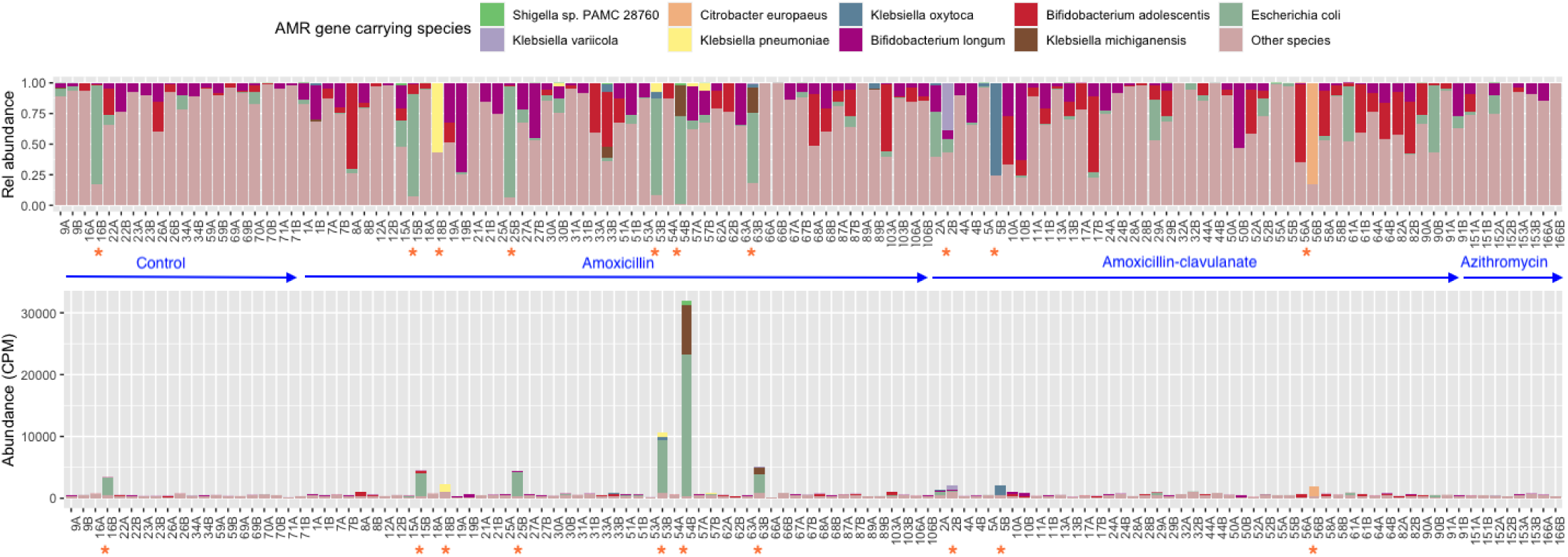
Composition and abundance of the AMR genes in the microbiome. The top plot shows the relative composition and the bottom plot shows the abundance in CPM. Both plots are colored by the source of species the AMR genes belong to. Each column is one sample. Sample IDs ending with ‘A’ and ‘B’ are samples before and after the treatment respectively. Subjects with visible increase in AMR are marked with “*”.

### Microbiome colonization resistance

The microbiome plays an important role in colonization resistance to prevent invasive species from colonizing the human gut. With deficient colonization resistance, humans have a higher risk of being infected by harmful AMR species, as seen in the children (subjects 16) in the control group. And certain antibiotics, such as amoxicillin and amoxicillin-clavulanate in this study, made the colonization resistance even weaker.

Here we were interested in finding the features in the baseline microbiome associated with the ability of colonization resistance. We divided the baseline samples into two groups: a vulnerable group experiencing visible AMR gene increase (subjects: 2, 5, 15, 16, 18, 25, 53, 54, 56, 63) and a resilient group for the remaining samples. The abundance of 5 genera, *Blautia, Ruminococcus, Faecalibacterium, Roseburia* and *Faecalitalea* in the resilient group were significantly higher than the vulnerable group (p < 0.001, FDR < 0.05, Wilcoxon rank sum test) (**Figure 5**). Those genera associated with resilient microbiome colonization resistance in the present study cohort are known short chain fatty acids (SCFAs) producers. Given the many well-known benefits of SCFAs produced by gut microbiome^29,30^, it is reasonable to speculate that SCFAs may play a role in modulating microbiome colonization resistance.

**Figure 5.**
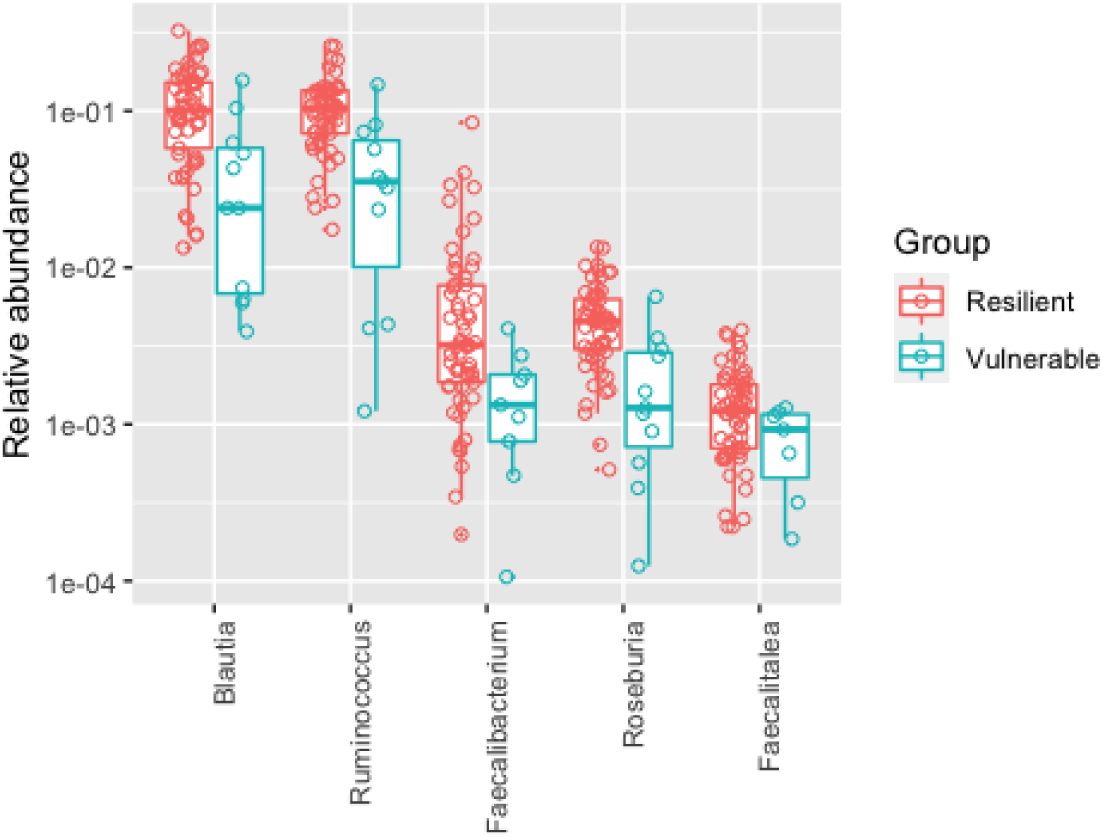
Significant protective features in baseline microbiome providing colonization resistance.

## Conclusion

In this study, we investigated how antibiotic treatment affects the intestinal microbiome and resistome, using a randomized controlled cohort of children. As it is not possible to investigate children in an experimental, prospective way and expose them to antibiotics without an indication for the treatment, we used non-severe acute otitis media as an excellent clinical model. This was possible in an ethically acceptable manner since current clinical guidelines suggest several alternative approaches to treat children with non-severe otitis media, including amoxicillin, amoxicillin-clavulanate, or watchful waiting without antibiotics.

We found that amoxicillin and amoxicillin-clavulanate didn’t have a significant impact on the total bacterial load, while azithromycin did cause the depletion of microbiome communities. Similar to what many previous studies showed, antibiotics treatment reduced the microbial diversity. We found that the resistome fluctuated over time. Even in the control group without any antibiotic treatment, there was a subject whose resistome increased rapidly due to colonization of AMR species. Antibiotic treatment by amoxicillin significantly increased this risk further, by allowing for invasive species, especially pathogenic species carrying AMR genes, to colonize the gut. Amoxicillin-clavulanate also increased this risk, but had a weaker effect than amoxicillin. Azithromycin, on the other hand, was able to sustain a more stable resistome. However, because our azithromycin group only had 4 children, this observation needs to be further confirmed. We also found that children lacking *Blautia, Ruminococcus, Faecalibacterium, Roseburia*, and *Faecalitalea* in the gut microbiome were more vulnerable to colonization by invasive AMR species. Here, we speculate that SCFAs produced by gut microbiome may play a role in modulating microbiome colonization resistance.

Besides the unique study cohort, we used a high-depth whole metagenomic sequencing approach to analyze the taxonomy, function and resistome of the microbiome. In addition, we also utilized flow cytometry-based approach to quantify absolute bacterial load in the samples. Our analyses were drawn from both relative abundance and absolute abundance of the microbial species in the samples.

Our findings are highly important with respect to public health and the control of AMR, as well as for clinical medicine, including primary care and decisions that are made on antibiotic treatment of children.

Certainly, this study has some limitations. First, our cohort recruitment was badly interrupted due to the COVID-19 world pandemic that reduced our sample size due to a rapid and surprising large reduction of acute otitis media incidence^31,32^. In particular, the azithromycin treatment group was too small. This was because only the children who were allergic to amoxicillin received azithromycin. Second, we only collected two samples per child before and after the treatment. So, our study only illustrated the short time effect of antibiotics treatment on microbiome and the resistome.

## Materials and methods

### Study cohort and sample collection

The participants were recruited in a randomized controlled trial (ClinicalTrials.gov ID NCT02935374, Eudra-CT: 2016-002927-27) designed to investigate the impact of antimicrobial therapy on gut microbiome in children with non-severe acute otitis media. The study population of 62 children was 50% male and 50% female, with 97.4% white and native Finnish (**Table S1**). Most children underwent testing for *Clostridium difficile* and antimicrobial resistant bacteria in the fecal samples before and after antibiotics according to the study protocol (**Table S1**). Children of 6 month to 6 years of age with non-severe otitis media were randomly allocated to three even-sized groups to receive either a 7 day dose of amoxicillin, amoxicillin-clavulanate, or no treatment. Four (4) children who were allergic to amoxicillin received azithromycin. Eleven (11) children, who were initially allocated to receive no treatment (Control group) but received antibiotics during the follow up visits, were finally included in antibiotics groups. So, the control, amoxicillin, amoxicillin-clavulanate and azithromycin each had 10, 25, 23 and 4 children respectively. The Regional Ethics Committee of the Northern Ostrobothnia Hospital District, Oulu University Hospital, Oulu, Finland, reviewed and approved the study plan (decision number EETTMK:82/2016). The clinical study was performed according to relevant regulations and guidelines. We enrolled only the children whose parents gave written informed consent.

Stool samples before the antibiotic treatment (Day 0) and 10 days after were collected using protocols in our previous studies ^11,33,34^ and stored at -80°C before shipping to the J. Craig Venter Institute (JCVI) on dry ice. After receiving the samples at the JCVI, samples were stored at -80°C until DNA extraction.

### Metagenomic sequencing

For DNA extraction, about 0.2 gram specimens were taken from each stool sample and 1 ml PBS were added to homogenize the stool materials. DNAs were extracted from fecal samples using the QIAamp PowerFecal Pro DNA Kit (Qiagen) following manufacturer protocols. Sequencing libraries were prepared from extracted DNA using the NEBNext® Ultra(tm) II kit (New England Biolabs). The libraries were sequenced on a NovaSeq 6000 instrument (Illumina) with a S2 flow cell following Xp v1.5 workflow for 300 cycles to yield 2×150 bp paired end (PE) reads.

On average, each sample was sequenced at a depth of 30.4 ± 7.8 million PE reads, or 9.1 ± 2.3 GB. After quality filtering (see next Metagenomic sequence analysis), 26.9 ± 7.2 million high quality PE reads (8.1 ± 2.1 GB) were used for the analysis (**Table S2**).

### Metagenomic sequence analysis

Raw reads were processed using Trimmomatic^35^ to remove low-quality bases and adapter sequences. High-quality paired end reads with both read longer than 100bp were mapped to the human reference genome (hg38) with BWA-MEM ^36^ and removed if they mapped concordantly with an alignment score of ≥60. We maintain a comprehensive microbiome reference genome database for mapping the metagenomic reads. This database was compiled and curated from NCBI Refseq genomes covering complete and draft bacteria, archaea, viruses, fungi and microbial eukaryotes species. Currently, the database contains 29,324 representative genomes. Taxonomy profiles were determined through reference genome mapping using Centrifuge^37^.

After initial mapping using Centrifuge, we first filtered out mapped genomes from contamination. The depth of coverage (DoC) (read length × number of mapped reads / genome length) and fraction of coverage (FoC) (number of bases covered by mapped reads / genome length) for each mapped genome was calculated using the alignment file provided by Centrifuge. Some genomes were filtered out using the following criteria. In sequencing, the observed number of times a base is sequenced follows a Poisson distribution^38^. Given the observed DoC calculated from the alignment file, the expected FoC is (1 - 1/(e^DoC^))^38^. If the observed FoC was smaller than 1/10 of expected value, then it suggested aligned reads were piled up at a small fraction on the genome rather than uniformly distributed along the genome, the genome was removed. Then the DoC for the remaining mapped genomes were normalized to relative abundance, so that the sum of relative abundances are added to 1.0 (100%). Taxonomy profiles at different ranks (phylum, genus, species) were calculated by adding relative abundances of all the mapped genomes within the taxa. Genomes with less than 10^−4^ abundance were not further considered in the analysis.

High-quality non-human reads were assembled into contigs using metaSPAdes ^39^. Contigs shorter than 250 bp were removed. The depth of coverage (DoC) for each contig, which is the total length of the reads assembled into this contig divided by the length of the contig, as reported by metaSPAdes, was saved. Protein-coding genes were predicted using Prodigal ^40^ from the contigs. Each gene inherits the DoC from the contig it was called. The DoCs were then converted into relative abundance as copy per million (CPM), which is the number of copies of a gene per million copies of total genes found in a sample (CPM_gene_ = DoC_gene_ / ∑_i = 1,n_ DoC_i_ × 1,000,000).

Protein-coding genes were annotated by comparing to an internally curated KEGG^41^ protein reference database using using cd-hit-2d (version 4.8.1)^42,43^ (-c 0.75 -n 5 -d 0 -g 1 -G 0 -aS 0.9 -A 60 -aL 0.25) to identify highly confident matches. Abundance of KEGG Ortholog (KO) clusters and pathways were calculated based on cd-hit-2d search. The abundance of a KO cluster (CPM) is the sum of CPMs of all genes that match this KO cluster in the cd-hit-2d search.

Protein-coding genes were processed using the Resistance Gene Identifier (RGI) in strict mode to predict the antibiotic resistance genes of each metagenomic sample using the CARD database ^27^. The abundance of an AMR family was converted to CPM, by transferring the CPM values from the genes.

### Bacteria load quantification

Bacterial cell counts in the stool samples were measured by flow cytometry. Concentrations of viable and dead bacterial cells were determined using a commercially available LIVE/DEAD(tm) BacLight(tm) Bacterial Viability and Counting Kit (ThermoFisher) following manufacturer protocols on a customized BD Fluorescence-activated Cell Sorting (FACS) Aria II flow cytometer. Homogenized stool specimens were first diluted 100,000 times. Stool samples were stained with membrane permeable SYTO 9 and membrane impermeable propidium iodide dyes and counted against a calibrated suspension of microspheres for accurate viability and sample volume measurements. Our custom Forward Scatter PMT equipped on the FACS Aria resolved events in the size range of 0.1 mm to 10 mm to encompass the requisite bacterial size ranges while allowing for discrimination of single bacteria from aggregates of 2 or more. Cell-counting was replicated three times for each sample and the mean was used in the analysis. In this study, the bacterial load values were represented as the number of total cells, live and dead, per gram of stool specimen.

With the known total bacteria load in the samples, the taxonomy abundance of mapped genomes were also converted to absolute abundance, in units of number of cells per gram of stool. When using absolute abundance, species with less than 10^6^ cells per gram of stool were not further considered in the analysis.

## Data availability

The raw metagenomic reads were deposited at NCBI and are available from SRA under bioproject access number PRJNA800433 from either https://www.ncbi.nlm.nih.gov/bioproject/PRJNA800433 or https://www.ncbi.nlm.nih.gov/sra/PRJNA800433

## Data Availability

The raw metagenomic reads were deposited at NCBI and are available from SRA under bioproject access number PRJNA800433.

https://www.ncbi.nlm.nih.gov/bioproject/PRJNA800433

## Acknowledgement

The work was supported by federal funds from the National Institute of Allergy and Infectious Diseases (NIAID), National Institutes of Health (NIH), Department of Health and Human Services under Award Number [1R21AI151730 to WL]. We thank the J Craig Venter Institute sequencing core for metagenomic sequencing.

## Supplementary materials

**Figure S1.**
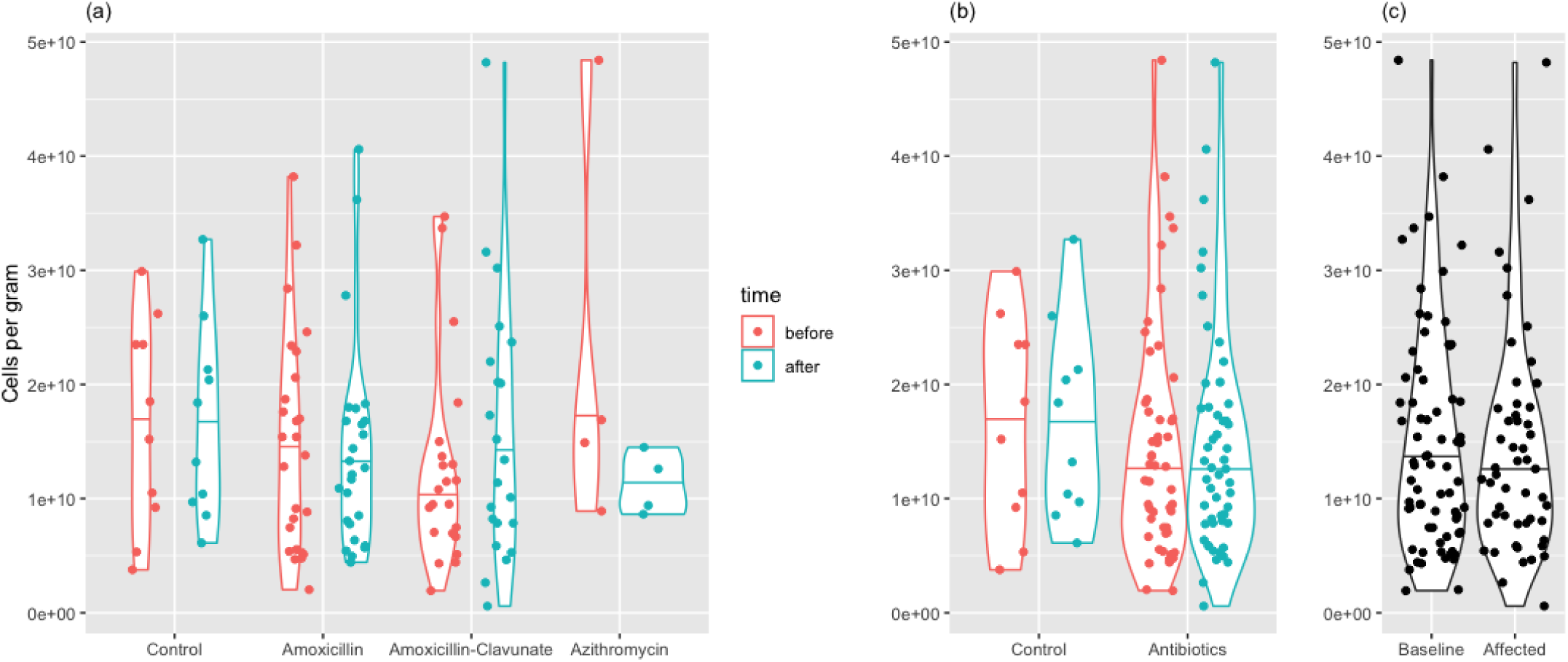
Bacterial cell count in fecal specimens before and after antibiotic treatments in the treatment and control groups (a), (b) and between the baseline and affected groups (c).

**Figure S2.**
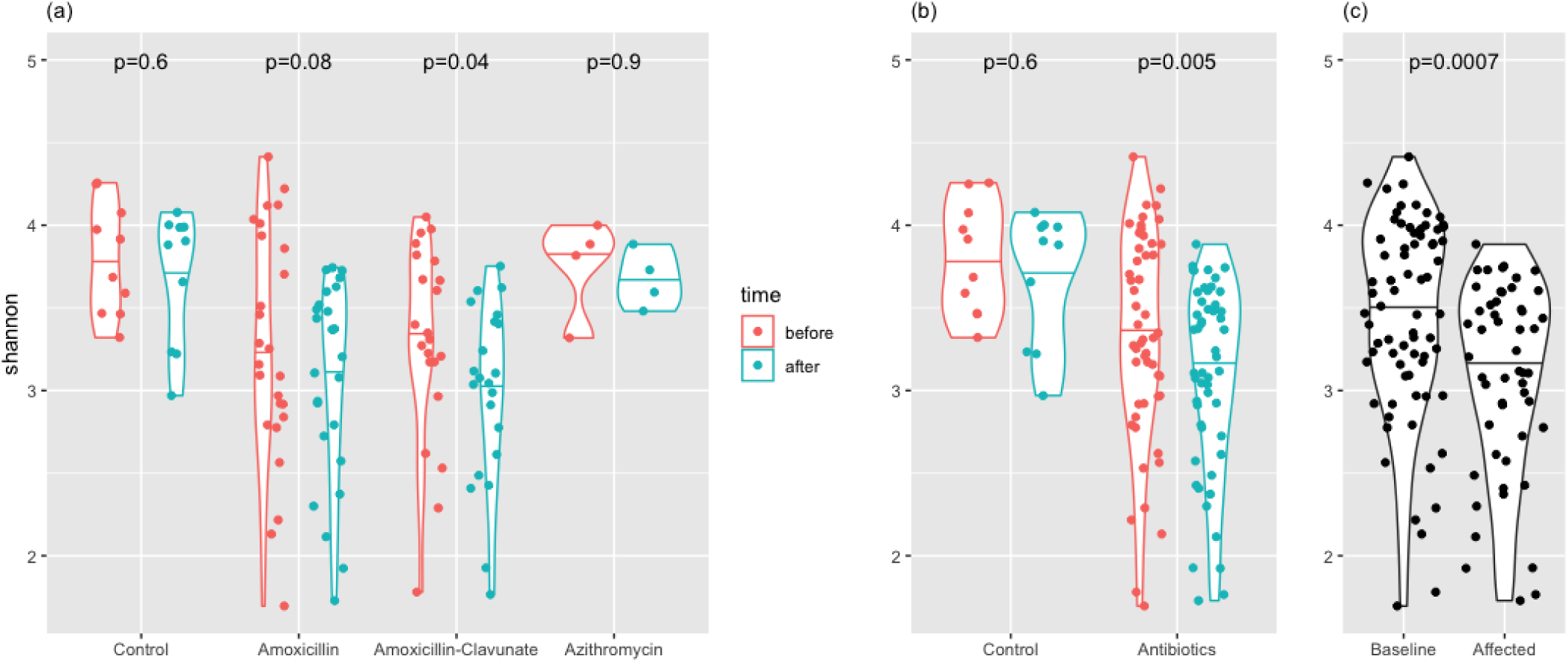
Shannon diversity before and after antibiotic treatments in treatment and control groups (a), (b) and between the baseline and affected groups (c).

**Figure S3.**
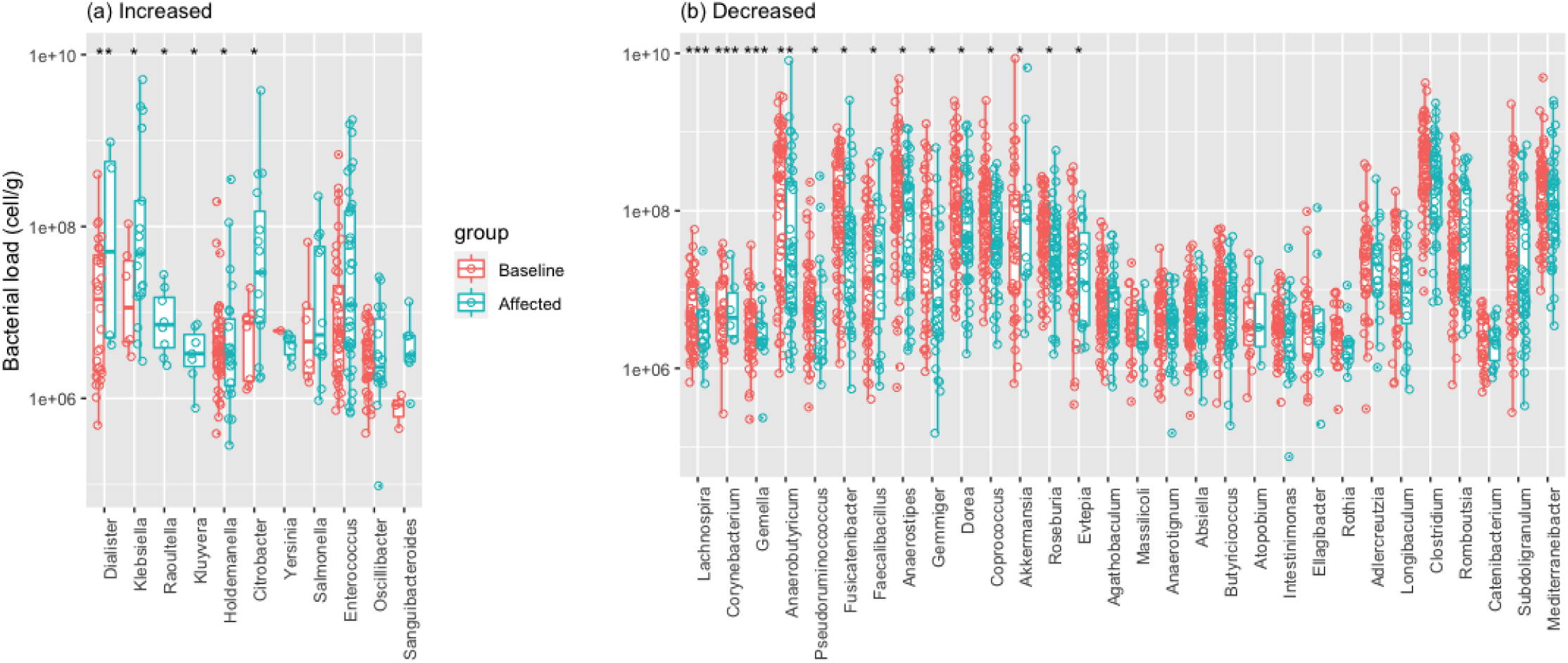
Genera impacted by antibiotic treatment. Similar to Figure 1, but this is based on the comparison between the baseline and the affected groups. Statistical significance annotation symbol: * P ≤ 0.05, ** P ≤ 0.01, *** P ≤ 0.001.

## Notes

### Competing Interest Statement

The authors have declared no competing interest.

### Clinical Trial

NCT02935374

### Author Declarations

The Regional Ethics Committee of the Northern Ostrobothnia Hospital District, Oulu University Hospital, Oulu, Finland, reviewed and approved the study.

